# Secondary analysis of the Game of Stones trial of text messages with financial incentives for men with obesity

**DOI:** 10.1101/2024.12.19.24319336

**Authors:** Stephan U Dombrowski, Pat Hoddinott, Lisa Macaulay, Catriona O’Dolan, James Swingler, Seonaidh Cotton, Alison Avenell, Abraham M Getaneh, Cindy Gray, Kate Hunt, Frank Kee, Alice MacLean, Michelle C McKinley, Claire Torrens, Katrina Turner, van der Pol Marjon, Graeme MacLennan

**Author notes:** **Contact info**: Stephan U. Dombrowski, PhD; 90 Mackay Drive, Fredericton, New Brunswick, Canada, E3B 5A3.

## Abstract

**Objective:** To explore whether socio-economic, health and behavioural characteristics moderate effectiveness of a text message intervention with or without financial incentives versus a control group, and to examine differences in exploratory outcomes.

**Methods:** Three-group randomized trial including 585 men with obesity comparing daily automated behavioural text messages for 12-months alongside financial incentives; text messages alone; or a waiting list control. Moderator analyses examined percent weight change after 12 months for 9 socio-economic and 11 health factors. Exploratory outcomes included: self-reported physical activity, sedentary behaviour, smoking and alcohol behaviours, engagement in 15 weight management strategies, and weight-management related confidence.

**Results:** No moderator effects were found by any factors for either comparison versus control. There were no differences between groups for health behaviours. The texts with incentives group had higher levels of engagement in six strategies including weight goals, food changes and self-weighing, and higher levels of confidence compared to the control group.

**Conclusion:** No evidence of differential intervention effectiveness was found across socio-economic, health or wellbeing status. The texts and financial incentives group showed greater engagement in weight management and favourable changes in weight management confidence compared to the control group.

## Introduction

Obesity is increasingly a worldwide problem and elevates the risk of adverse health conditions for individuals carrying excess fat on their body(1). Although approximately 26% of UK and 43% of US adult men are estimated to be living with obesity(2, 3), evidence suggests that men are less likely than women to engage in weight management interventions, programmes and services(4). Moreover, evidence gaps remain for engagement and effectiveness of weight management interventions in men living with obesity, particularly in those who also report low socio-economic status(5, 6).

Behavioural weight management interventions seeking volitional changes in eating behaviours and physical activity remain a cornerstone of accessible low-risk obesity treatment. Equitable, inclusive, low-burden and scalable Interventions are required to address health inequalities associated with obesity and engage underserved populations. It is important for behavioural interventions to avoid intervention-generated escalation of inequalities and contribute towards improving obesity-related health at a population level(7).

The Game of Stones trial randomized 585 men with obesity to behavioural text messages with financial incentives, text messages alone or a waiting list control group(8). Findings showed a 4.8% weight loss at 12 months in participants who received text messages with financial incentives, which was significantly different from the control group who lost 1.3% of their baseline weight(9). The text messages alone group lost 2.7% which was not significantly different to the control group. Game of Stones is a remotely delivered low-burden intervention with direct in-person contact limited to four brief weight assessments over 12 months. Whilst the trial results overall are positive, the intervention requires further examination to ensure that it does not disproportionately affect vulnerable subgroups, such as those disadvantaged by socio-economic circumstances or health; and examine effectiveness in relation to exploratory outcomes such as behavioural changes, engagement in weight management strategies and psychological variables.

This secondary analysis aims to explore whether baseline socio-economic, health and wellbeing characteristics moderate effectiveness of the primary outcome of percent weight change at 12 months for men with obesity randomized to a text message intervention with, or without, financial incentives versus a control group, and to examine differences in exploratory outcomes.

## Methods

### Intervention

The Game of Stones trial was a three-arm parallel group, assessor blinded randomized clinical trial conducted between July 2021 to July 2023 in three UK areas: Belfast, Bristol and Glasgow(8, 9). Men were invited through family practices, community information and social media targeting disadvantaged areas. Overall, 585 men were recruited with a body mass index ≥30kg/m^2^.

The three study arms were: i) daily automated behaviour-focused text messages designed to support weight management for 12 months alongside loss-framed incentives in which money was ‘lost’ from an initial endowment of $490 (£400) by not meeting verified weight loss targets (5% at 3 months, 10% at 6 months and maintaining 10% weight loss at 12 months), in comparison with baseline weight; ii) text messages (as described in i) above) alone; or iii) a 12-month waiting list for three months of text messages. All groups received access to a website containing evidence-based weight management information and a pedometer at baseline. Intervention groups also received localised webpages signposting to services and self-monitoring web pages.

The study received ethical approval from the North of Scotland Research Ethics Committee 2 [20/NS/0141] and the protocol has been published(8).

### Outcomes and assessments

Outcomes and assessments were based on the Game of Stones feasibility trial(10) which included extensive public, patient and stakeholder involvement to assess acceptability and burden of data collection tools informed by guidance on outcomes in weight management trials (STAR-LIGHT(11)), PROGRESS-Plus characteristics(12) and CONSORT equity reporting guidance(13). The study balanced potential academic and participants’ benefits and harms of data collection (14).

Baseline data were collected before randomisation and used previously piloted(10) and validated measures, where available. No consensus on the most appropriate measures to evaluate behavioural weight management interventions in men with obesity currently exists. Outcomes were selected considering the different study recruitment routes of community and primary care. Participants included both younger men who were not engaging in health services and older men with multiple long-term conditions and disability.

Pre-specified subgroup analyses for moderators of the primary outcome of percent weight change at 12 months from baseline were undertaken within three categories: i) socio-economic factors; ii) health and wellbeing status, and iii) recruitment route.

#### Socio-economic factors

The assessments of level of disadvantage included use of the Index of Multiple Deprivation (IMD) which is a measure of relative deprivation based on UK postcode address where participants live, drawing on variables such as income, education and crime rates. The IMD can be used to divide the population into five deprivation categories which, for the current analysis, were aggregated into the two more deprived categories compared to the three more affluent categories. Data from England, Scotland and Northern Ireland were classified as per the country-specific methodology for allocation of IMD subgroup classification(12, 15).

Guidance published by the UK Office for National Statistics(16) was used to harmonise and score key individual level variables including (participant) education (university degree level or above versus other qualification versus no qualification), living status (living alone versus living with others) and relationship status (single versus married/in a partnership). The harmonised guidance from the Scottish Government(17) was used to assess working status (in paid work/self-employed versus unpaid).

Perceived wealth was assessed using three items)(18) (e.g. “I feel that I have enough money”) scored from 0 (strongly agree) to 100 (strongly disagree) and dichotomised into low (≤50) and high (≥51). The perceived wealth measures were unintentionally reverse scored, with lower scores indicating higher perceived wealth, unlike the original measure where higher scores indicate higher perceived wealth.

Financial strain was assessed using one item based on French (2017) (19) (‘How well would you say you yourself are managing financially these days?’) with five possible response options, dichotomised into easier (‘living comfortably’, ‘doing alright’ and ‘just about getting by’) versus harder (‘finding it quite difficult’ and ‘finding it very difficult’).

#### Health and wellbeing factors

Quality of life was assessed using the EQ-5D-5L overall utility score (dichotomised into high [above 0.4005] versus low [below at 0.4005]) and EQ-5D-5L Anxiety and Depression dimension (dichotomised into low [1-3] versus high [4-5])(20).

Mental wellbeing was assessed using the Warwick-Edinburgh Mental Well-being Scale (WEMWBS)(21) consisting of 14 items (e.g. “I’ve been feeling optimistic about the future”) scored from 1 (none of the time) to 5 (all of the time) dichotomised into low (≤40) versus high ≥41).

Mental health was assessed using four items of the Patient Health Questionnaire-4 (PHQ-4)(22), consisting of an anxiety subscale (GAD-2, 2 items) and a depression subscale (PHQ-2, 2 items). Items were scored from 0 (not at all) to 3 (nearly every day) and summed and dichotomised into high (≥3 for GAD-2 or PHQ-2) versus low (≤2 for GAD-2 or PHQ-2).

Perceived weight-related stigma was assessed using the Weight Self-Stigma Questionnaire (WSSQ) (23) consisting of 12 items (e.g. “I feel guilty because of my weight problems”) scored from 0 (completely disagree) to 5 (completely agree) and dichotomised into high (≥42) versus low (≤41).

Co-morbidities were assessed with the item “Has a doctor ever told you that you have/had…?” followed by the response options ‘a stroke (including mini-stroke)’, ‘high blood pressure’, ‘a heart condition such as angina or atrial fibrillation’, ‘diabetes’, ‘cancer’, ‘arthritis’, and ‘a mental health condition’ (dichotomised into yes for those reporting at least one co-morbidity versus no for those reporting none). The presence of multiple long-term conditions (MLTC) was defined as the co-existence of two or more co-morbidities. In addition, a self-reported mental health condition (yes versus no) and diabetes (yes versus no) were analysed separately in subgroup analyses.

A variable labelled ‘Possible Latent Mental Health Condition’ was defined for men who did not self-report a mental health condition but whose scores on at least one of the PHQ-4, EQ-5D-5L-AD, WEMWBS or WSSQ exceeded a threshold suggesting a possible undetected mental health condition (see above for scoring details).

Self-reported disability was assessed with the two items based on Office for National Statistics definitions(24): “Do you have any physical or mental health conditions or illnesses lasting or expected to last 12 months or more?” and “Do any of your conditions or illnesses reduce your ability to carry-out day-to-day activities?”. Those answering yes to both were defined as having a disability.

Alcohol consumption was measured using a single question (“During the last month, how many days did you usually have any kind of drink containing alcohol?”) with eight possible response options ranging from ‘Never’ to ‘Everyday’ (dichotomised into drinking every day versus not every day).

#### Recruitment

Participants were categorised according to the route of recruitment (community-based versus via general practice).

#### Secondary exploratory outcomes

Physical activity and sedentary behaviour were assessed from the self-reported number of days of vigorous and moderate physical activity and time spent sitting respectively, using the International Physical Activity Questionnaire(25).

Smoking status was measured with one item (“Do you currently smoke or have you ever smoked?”) with response options ‘Yes, I currently smoke every day’, ‘Yes, I currently smoke, but not every day’, ‘Yes, I used to smoke but have quit’, and ‘No, I have never smoked’.

Self-monitoring of activity and weight were assessed with one item respectively (“How often do you monitor your steps?”, “How often do you keep track of your weight by weighing yourself”?) with six response options ranging from ‘Never’ to ‘Everyday’.

Weight management strategies were assessed with the item: “Which of these strategies have you used in the last 12 months to lose weight?”. Participants were provided with 13 response options (e.g. “Had a weight goal to work towards”) based on evidence of effective strategies for weight management(26).

Confidence in ability to lose weight and confidence in ability to maintain weight loss long-term were each assessed with a single item (“How confident are you in your ability to lose weight?”, “How confident are you in your ability to keep lost weight off in the long term?”), with responses on a 7 point scale ranging from 1 (not confident) to 7 (very confident).

#### Sample size calculation

The sample size calculation for this trial was for the primary outcome of percentage weight change from baseline and 12 months(9).

### Analysis

The primary outcome subgroup modelling used linear regression adjusted for the recruitment areas (Belfast, Bristol, Glasgow) and recruitment route (family practice or community), treatment group, the subgroup of interest and a treatment-by-subgroup interaction term. Confidence intervals are presented at 99.5% to reflect the number of subgroups tested and the exploratory nature of analysis, equivalent to a stringent level of evidence required for significance of p < 0.005. Results are summarised as Forest plots of within-subgroup treatment effects and the interaction term testing the moderating effect of the subgroup.

Subgroup analyses are split into confirmatory and exploratory. Confirmatory subgroup analyses (as pre-specified in the statistical analysis plan) included obesity-related comorbidity (present versus absent) and diabetes (present versus absent). The confirmatory subgroup analyses are based on hypothesized directions of effect modification of the interventions informed by the weight-loss literature (27). Weight loss and/or weight loss maintenance are part of disease management for many obesity-related co-morbidities, e.g. diabetes, cardio-vascular disease. All other pre-specified subgroup analyses were designated as exploratory.

Secondary exploratory outcomes were analysed using a generalized linear model suitable for the outcome distribution, adjusting for recruitment centre, recruitment route and the baseline measure of the outcome if measured. Confidence intervals for all secondary outcomes are presented at the 97.5% for all secondary outcomes.

## Results

A total of 585 participants were randomised to text messaging with financial incentives group (n=196), text messaging alone group (n=194), or the waiting list control group (n=195), and 73% of participants (n=426) provided weight data at 12 months.

Key baseline characteristics are reported in Table 1. Intervention groups were comparable across trial groups – for information on all assessed baseline characteristics see (9). Participants had a mean BMI of 37.7kg/m^2^ (SD, 5.7) and a mean age of 50.7 (SD, 13.3) years. Most were of white ethnicity (93%), married/ living with a partner (62%), and reported one or more co-morbidities (71%), including 18% of participants overall reporting diabetes.

**Table 1:**
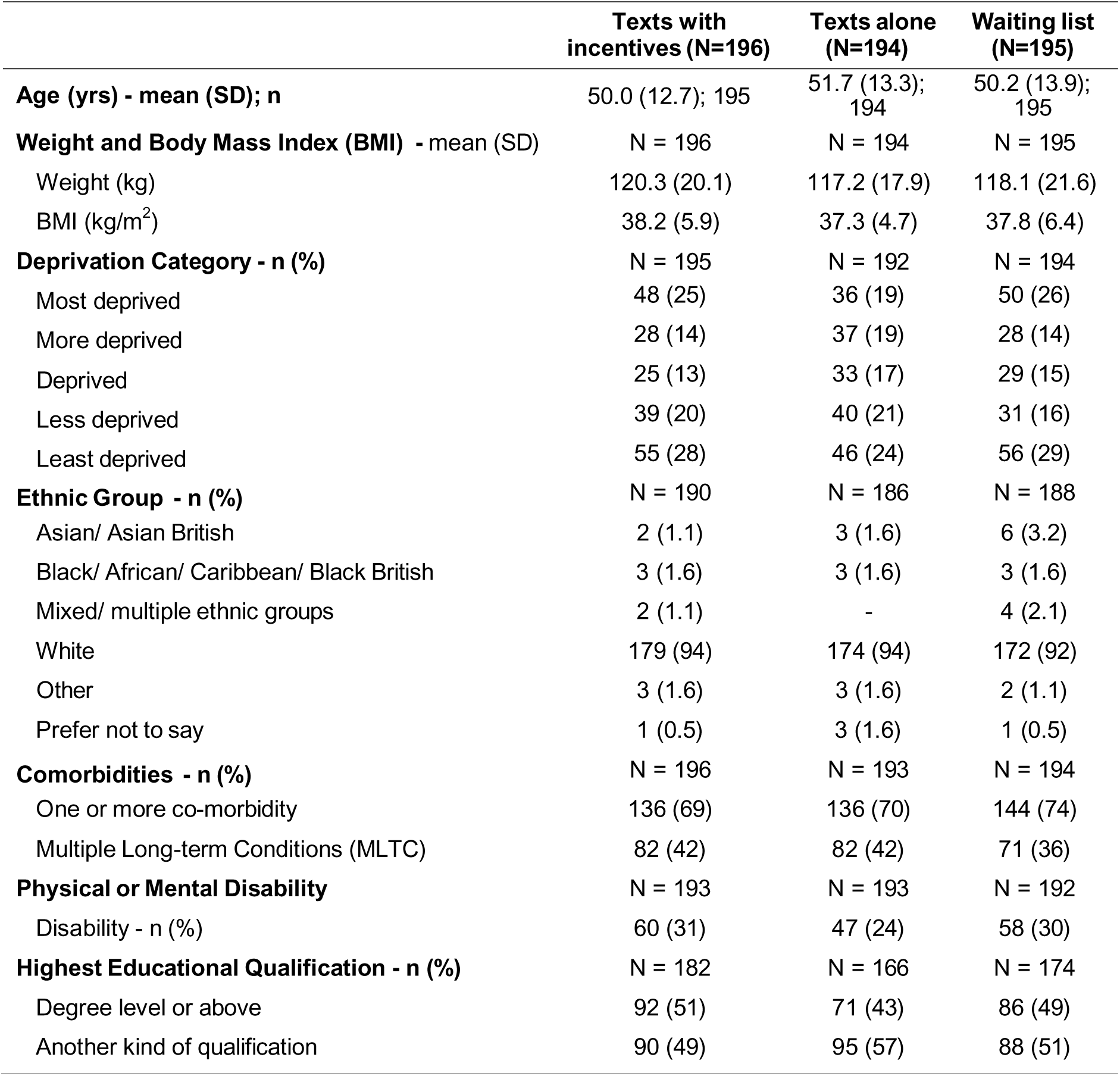
Baseline characteristics by treatment allocation.

|  | **Texts with**<br>**incentives (N=196)** | **Texts alone**<br>**(N=194)** | **Waiting list**<br>**(N=195)** |
| --- | --- | --- | --- |
| **Age (yrs) - mean (SD); n** | 50.0 (12.7); 195 | 51.7 (13.3);<br>194 | 50.2 (13.9);<br>195 |
| **Weight and Body Mass Index (BMI) - mean (SD)** | N = 196 | N = 194 | N = 195 |
| Weight (kg) | 120.3 (20.1) | 117.2 (17.9) | 118.1 (21.6) |
| BMI (kg/m<sup>2</sup>) | 38.2 (5.9) | 37.3 (4.7) | 37.8 (6.4) |
| **Deprivation Category - n (%)** | N = 195 | N = 192 | N = 194 |
| Most deprived | 48 (25) | 36 (19) | 50 (26) |
| More deprived | 28 (14) | 37 (19) | 28 (14) |
| Deprived | 25 (13) | 33 (17) | 29 (15) |
| Less deprived | 39 (20) | 40 (21) | 31 (16) |
| Least deprived | 55 (28) | 46 (24) | 56 (29) |
| **Ethnic Group - n (%)** | N = 190 | N = 186 | N = 188 |
| Asian/ Asian British | 2 (1.1) | 3 (1.6) | 6 (3.2) |
| Black/ African/ Caribbean/ Black British | 3 (1.6) | 3 (1.6) | 3 (1.6) |
| Mixed/ multiple ethnic groups | 2 (1.1) | - | 4 (2.1) |
| White | 179 (94) | 174 (94) | 172 (92) |
| Other | 3 (1.6) | 3 (1.6) | 2 (1.1) |
| Prefer not to say | 1 (0.5) | 3 (1.6) | 1 (0.5) |
| **Comorbidities - n (%)** | N = 196 | N = 193 | N = 194 |
| One or more co-morbidity | 136 (69) | 136 (70) | 144 (74) |
| Multiple Long-term Conditions (MLTC) | 82 (42) | 82 (42) | 71 (36) |
| **Physical or Mental Disability** | N = 193 | N = 193 | N = 192 |
| Disability - n (%) | 60 (31) | 47 (24) | 58 (30) |
| **Highest Educational Qualification - n (%)** | N = 182 | N = 166 | N = 174 |
| Degree level or above | 92 (51) | 71 (43) | 86 (49) |
| Another kind of qualification | 90 (49) | 95 (57) | 88 (51) |

The main results have been published previously(9). The overall mean (SD) percent weight change was −4.8% (6.1%) for the financial incentives group, −2.7% (6.3%) for the text messaging group, and −1.3% (5.5%) for the control group. At the 12-month follow-up, the text messaging with incentives group had significantly greater weight loss (mean difference in percentage change from baseline, −3.2%;97.5% CI, −4.6 to −1.9; P < .001), and the text messaging alone group did not have significantly greater weight loss (mean difference in percentage change from baseline, −.4%; 97.5%CI, −2.9% to 0.0; P = .05, compared to the control group.

### Moderator analyses

Confirmatory subgroup analyses found no evidence for an interaction for the presence of a co-morbidity or diabetes for either the texts with incentives compared to the control group, or the texts alone group compared to the control group (*p-values for interactions* ≥.19, Table 2, Figures 1 and 2).

**Figure 1:**
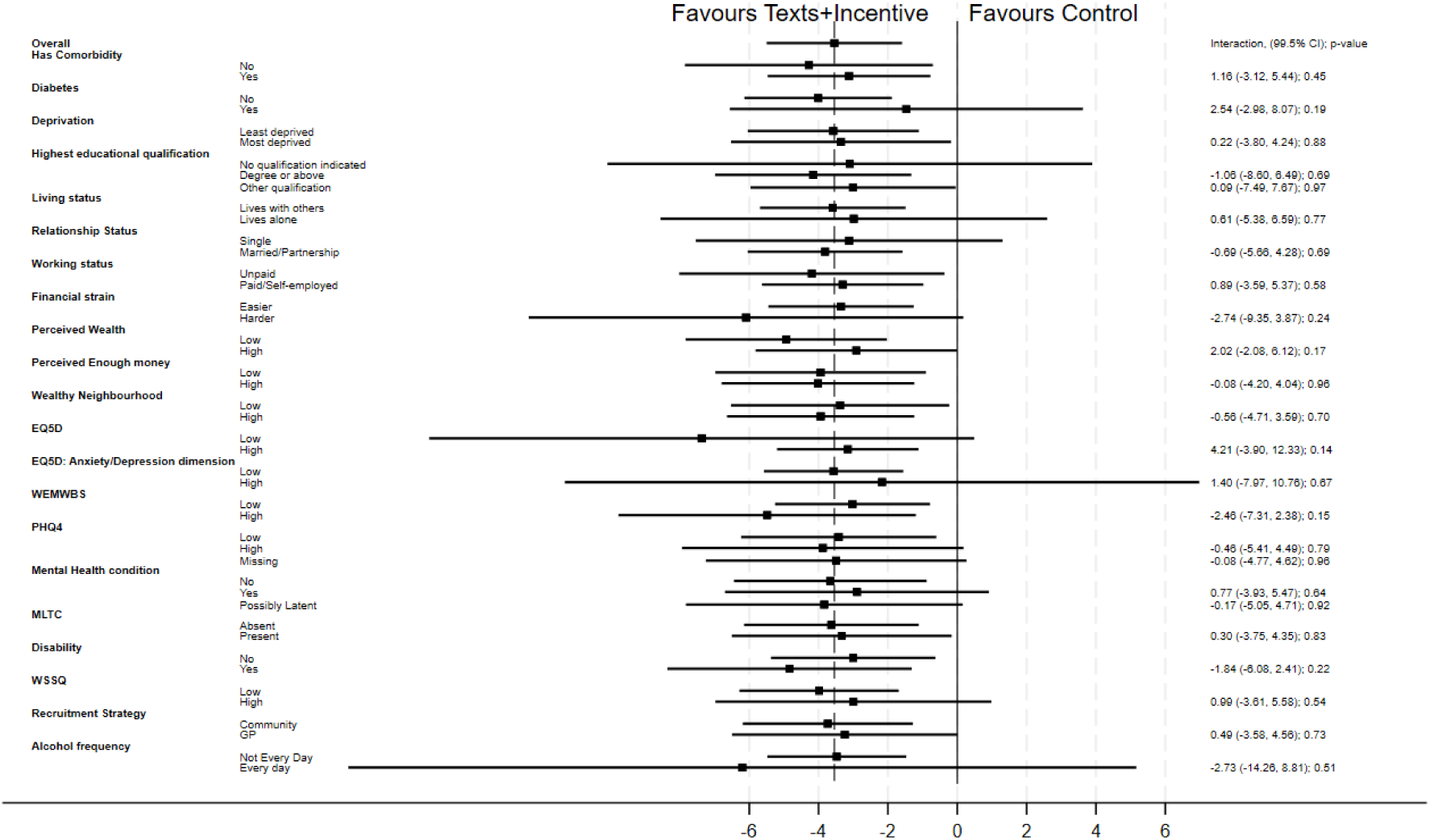
Forest plot for subgroup analysis comparing percent weight loss (99.5% confidence intervals) at 12 months from baseline between the texts with incentives group compared to the control group.

**Figure 2:**
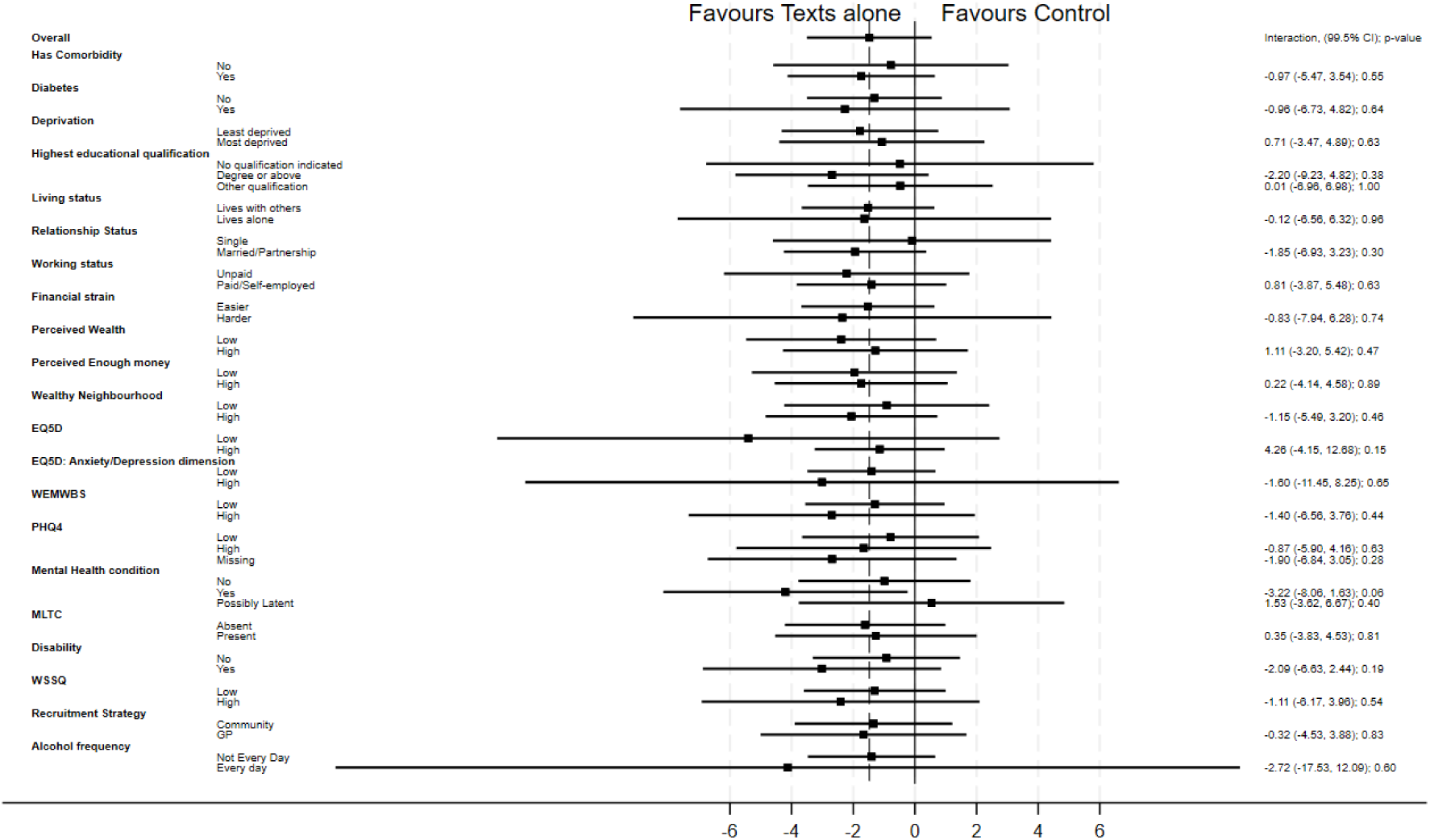
Forest plot for subgroup analysis comparing percent weight loss (99.5% confidence intervals) at 12 months from baseline between the texts alone group compared to the control group.

**Table 2:** Subgroup analyses for percent weight change at 12 months from baseline.

| Analysis type<br>Subgroup<br>category* | Texts with Incentives<br>(n=146) | Texts alone<br>(n=128) | Control<br>(n=152) | Texts with Incentives<br>versus Control<br>Interaction Effect:<br>Mean Difference<br>(99.5% CI); p value | Texts alone versus<br>Control<br>Interaction Effect: Mean<br>Difference (99.5% CI); p<br>value |
| --- | --- | --- | --- | --- | --- |
| **Confirmatory analyses – mean % weight change, (SD), n** |  |  |  |  |  |
| **Has Comorbidity** |  |  |  |  |  |
| No | -5.8 (5.3); 46 | -2.4 (6.6); 36 | -1.7 (6.4); 43 |  |  |
| Yes | -4.3 (6.5); 99 | -2.9 (6.2); 92 | -1.1 (5.1); 109 | 1.16 (-3.12, 5.44); 0.45 | -0.97 (-5.47, 3.54); 0.55 |
| **Has Diabetes** |  |  |  |  |  |
| No | -5.3 (6.0); 117 | -2.6 (6.5); 106 | -1.3 (5.7); 134 |  |  |
| Yes | -2.8 (6.7); 28 | -3.5 (5.3); 22 | -1.3 (3.8); 18 | 2.54 (-2.98, 8.07); 0.19 | -0.96 (-6.73, 4.82); 0.64 |
| **Exploratory Analyses – mean % weight change, (SD), n** |  |  |  |  |  |
| *Socio-economic Factors* |  |  |  |  |  |
| **Deprivation Category** |  |  |  |  |  |
| Less deprived | -4.3 (5.9); 90 | -2.4 (6.9); 80 | -0.7 (5.0); 95 |  |  |
| More deprived | -5.6 (6.6); 56 | -3.4 (5.0); 47 | -2.3 (6.1); 56 | 0.22 (-3.80, 4.24); 0.88 | 0.71 (-3.47, 4.89); 0.63 |
| **Highest Educational Qualification** |  |  |  |  |  |
| No qualification<br>indicated | -5.6 (7.3); 10 | -3.0 (5.3); 15 | -2.6 (4.1); 14 |  |  |
| Degree or above | -4.6 (5.8); 70 | -3.0 (6.8); 49 | -0.4 (5.3); 73 | -1.06 (-8.60, 6.49);<br>0.69 | -2.20 (-9.23, 4.82); 0.38 |
| Other qualification | -4.9 (6.4); 66 | -2.5 (6.1); 64 | -2.0 (5.8); 65 | 0.09 (-7.49, 7.67); 0.97 | 0.01 (-6.96, 6.98); 1.00 |
| **Living Status** |  |  |  |  |  |
| Lives with others | -4.6 (6.2); 127 | -2.5 (6.4); 115 | -1.0 (5.4); 133 |  |  |
| Lives alone | -5.8 (5.8); 18 | -4.5 (4.8); 13 | -2.9 (5.6); 19 | 0.61 (-5.38, 6.59); 0.77 | -0.12 (-6.56, 6.32); 0.96 |
| **Relationship Status** |  |  |  |  |  |
| Single | -5.0 (7.0); 28 | -1.8 (4.0); 26 | -1.9 (6.0); 31 |  |  |
| Married/Partnership | -4.8 (5.9); 116 | -3.0 (6.8); 101 | -1.0 (5.3); 116 | -0.69 (-5.66, 4.28);<br>0.69 | -1.85 (-6.93, 3.23); 0.30 |
| **Working Status** |  |  |  |  |  |
| Not in paid employment | -6.0 (6.5); 40 | -4.0 (5.5); 34 | -1.7 (4.9); 38 |  |  |
| Paid/Self-employed | -4.3 (5.9); 103 | -2.4 (6.6); 89 | -1.1 (5.7); 109 | 0.89 (-3.59, 5.37); 0.58 | 0.81 (-3.87, 5.48); 0.63 |
| **Financial Strain** |  |  |  |  |  |
| Easier | -4.4 (6.0); 127 | -2.6 (6.3); 114 | -1.1 (5.3); 131 |  |  |
| Harder | -8.3 (6.6); 15 | -4.5 (6.8); 11 | -2.1 (7.2); 14 | -2.74 (-9.35, 3.87); 0.24 | -0.83 (-7.94, 6.28); 0.74 |
| **Perceived Wealth** |  |  |  |  |  |
| Low | -5.4 (6.2); 77 | -2.8 (5.4); 61 | -0.5 (4.7); 64 |  |  |
| High | -4.3 (6.1); 64 | -2.7 (7.5); 57 | -1.4 (6.1); 76 | 2.02 (-2.08, 6.12); 0.17 | 1.11 (-3.20, 5.42); 0.47 |
| **Perceived Enough Money** |  |  |  |  |  |
| Low | -4.9 (6.7); 71 | -2.9 (6.0); 50 | -1.0 (4.6); 58 |  |  |
| High | -4.9 (5.7); 70 | -2.7 (6.8); 67 | -0.9 (5.9); 83 | -0.08 (-4.20, 4.04); 0.96 | 0.22 (-4.14, 4.58); 0.89 |
| **Perceived Wealth Compared to Neighbourhood** |  |  |  |  |  |
| Low | -4.9 (6.4); 66 | -2.5 (4.9); 52 | -1.6 (4.5); 55 |  |  |
| High | -4.8 (6.0); 75 | -2.9 (7.4); 68 | -0.9 (6.2); 88 | -0.56 (-4.71, 3.59); 0.70 | -1.15 (-5.49, 3.20); 0.46 |
| *Health and Wellbeing factors* |  |  |  |  |  |
| **EQ-5D-5L** |  |  |  |  |  |
| Low | -7.3 (8.6); 14 | -5.3 (7.4); 11 | 0.2 (6.4); 7 |  |  |
| High | -4.5 (5.8); 131 | -2.5 (6.1); 117 | -1.4 (5.5); 142 | 4.21 (-3.90, 12.33); 0.14 | 4.26 (-4.15, 12.68); 0.15 |
| **EQ-5D: Anxiety/Depression dimension** |  |  |  |  |  |
| Low | -4.9 (6.1); 139 | -2.7 (6.2); 123 | -1.3 (5.4); 144 |  |  |
| High | -3.1 (8.4); 6 | -3.6 (9.1); 5 | -0.7 (6.8); 8 | 1.40 (-7.97, 10.76); 0.67 | -1.60 (-11.45, 8.25); 0.65 |
| **WEMWBS** |  |  |  |  |  |
| Low | -4.4 (6.1); 107 | -2.6 (6.3); 103 | -1.4 (5.7); 121 |  |  |
| High | -6.1 (6.1); 35 | -3.2 (6.2); 25 | -0.4 (4.2); 28 | -2.46 (-7.31, 2.38); 0.15 | -1.40 (-6.56, 3.76); 0.44 |

| PHQ-4 |  |  |  |  |  |
| --- | --- | --- | --- | --- | --- |
| Low | -4.8 (6.2); 67 | -2.2 (6.4); 62 | -1.5 (5.3); 80 |  |  |
| High | -4.9 (6.3); 37 | -2.6 (6.1); 34 | -0.9 (5.5); 33 | -0.46 (-5.41, 4.49);<br>0.79 | -0.87 (-5.90, 4.16); 0.63 |
| Missing* | -4.7 (6.0); 42 | -3.9 (6.3); 32 | -1.2 (5.8); 39 | -0.08 (-4.77, 4.62);<br>0.96 | -1.90 (-6.84, 3.05); 0.28 |
| Mental Health Condition |  |  |  |  |  |
| No | -5.2 (5.9); 68 | -2.5 (4.1); 67 | -1.6 (5.2); 79 |  |  |
| Yes | -3.2 (6.6); 38 | -4.4 (8.0); 33 | -0.3 (6.0); 40 | 0.77 (-3.93, 5.47); 0.64 | -3.22 (-8.06, 1.63); 0.06 |
| Possibly Latent | -5.7 (5.9); 39 | -1.3 (7.8); 28 | -1.7 (5.3); 33 | -0.17 (-5.05, 4.71);<br>0.92 | 1.53 (-3.62, 6.67); 0.40 |
| Multiple Long-term Condition (MLTC) |  |  |  |  |  |
| Absent | -4.8 (6.1); 82 | -2.8 (7.2); 73 | -1.2 (5.8); 100 |  |  |
| Present | -4.8 (6.2); 63 | -2.7 (4.9); 55 | -1.4 (4.7); 52 | 0.30 (-3.75, 4.35); 0.83 | 0.35 (-3.83, 4.53); 0.81 |
| Disability |  |  |  |  |  |
| No | -4.6 (6.0); 98 | -2.5 (6.4); 95 | -1.6 (5.4); 105 |  |  |
| Yes | -5.3 (6.6); 46 | -3.4 (5.8); 33 | -0.4 (5.5); 46 | -1.84 (-6.08, 2.41);<br>0.22 | -2.09 (-6.63, 2.44); 0.19 |
| Weight Stigma (WSSQ) |  |  |  |  |  |
| Low | -5.3 (6.3); 101 | -2.6 (4.2); 102 | -1.3 (5.2); 116 |  |  |
| High | -4.0 (5.7); 42 | -3.4 (11.6); 25 | -0.9 (6.3); 32 | 0.99 (-3.61, 5.58); 0.54 | -1.11 (-6.17, 3.96); 0.54 |
| Alcohol Frequency |  |  |  |  |  |
| Not Every Day | -4.8 (6.2); 140 | -2.7 (6.3); 125 | -1.3 (5.5); 146 |  |  |
| Every day | -4.6 (4.6); 5 | -2.1 (1.3); 2 | 1.9 (2.5); 4 | -2.73 (-14.26, 8.81);<br>0.51 | -2.72 (-17.53, 12.09);<br>0.60 |
| Recruitment route |  |  |  |  |  |
| Recruitment |  |  |  |  |  |
| Community | -5.0 (6.0); 88 | -2.5 (6.2); 76 | -1.2 (6.0); 102 |  |  |
| GP | -4.6 (6.4); 58 | -3.0 (6.4); 52 | -1.4 (4.1); 50 | 0.49 (-3.58, 4.56); 0.73 | -0.32 (-4.53, 3.88); 0.83 |

Note: \* a missing subgroup was created if the missing element of a variable exceeded 10% of the total responses, subgroup analyses for social weight loss are displayed in Online Appendix 2.
CI = Confidence Interval, EQ-5D-5L = EuroQol-5 Dimension 5 Level scale, GP = General practice, n= Number, PHQ-4 = Patient Health Questionnaire-4, SD = Standard Deviation, WEMWBS = Warwick-Edinburgh Mental Well-being Scale, WSSQ = Weight Self-Stigma Questionnaire.

Exploratory subgroup analyses for socioeconomic factors found no evidence for an interaction for deprivation category, education, living status, relationship status, working status, financial strain, perceived wealth, perceived enough money, and perceived neighbourhood wealth for either the texts and incentives compared to the control group, or the texts alone group compared to the control group (*p-values for interactions* ≥.02, Table 2).^1^

Exploratory subgroup analyses for health and wellbeing status found no evidence for an interaction for overall quality of life (EQ-5D-5L), Anxiety/Depression (EQ-5D dimensions), mental wellbeing (WEMWBS), mental health (PHQ-4), self-reported mental health condition, multiple long-term condition, disability status, weight stigma, and alcohol consumption for either the texts with incentives compared to the control group, or the texts alone group compared to the control group (*p-values for interactions* ≥.06, Table 2).^2^

Exploratory subgroup analyses for recruitment route found no evidence for an interaction for either the texts with incentives compared to the control group, or the texts alone group compared to the control group (*p-value for interactions* ≥0.73, Table 2).

## Secondary outcomes

### Health behaviours

There were no statistically significant differences in self-reported number of days of vigorous and moderate physical activity or time spent sedentary between either the texts with incentives, or the texts alone groups compared to the control group (Table 3). Moreover, no statistically significant differences were found for alcohol consumption and smoking status between either intervention group compared to the control group.

**Table 3:**
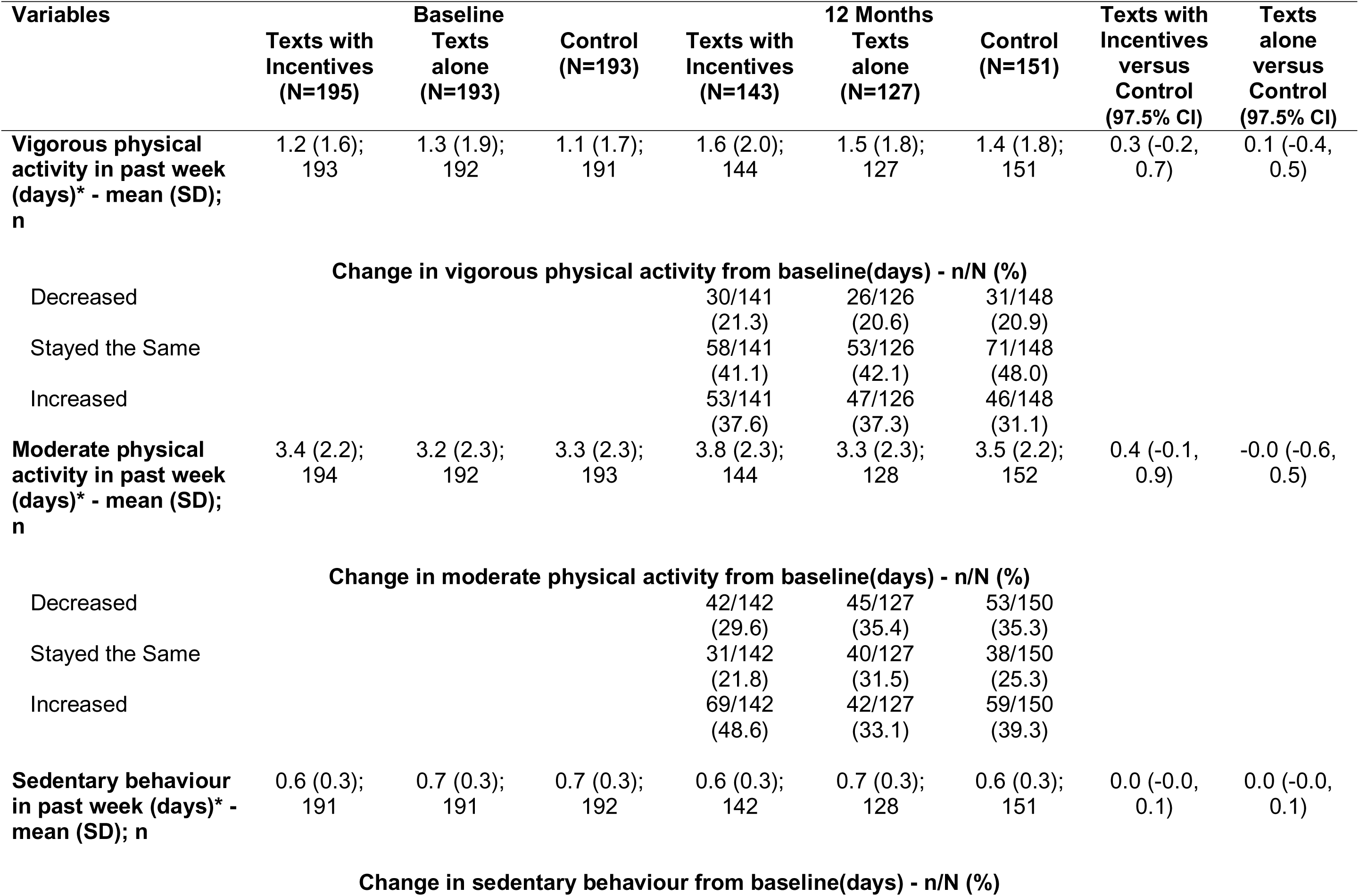
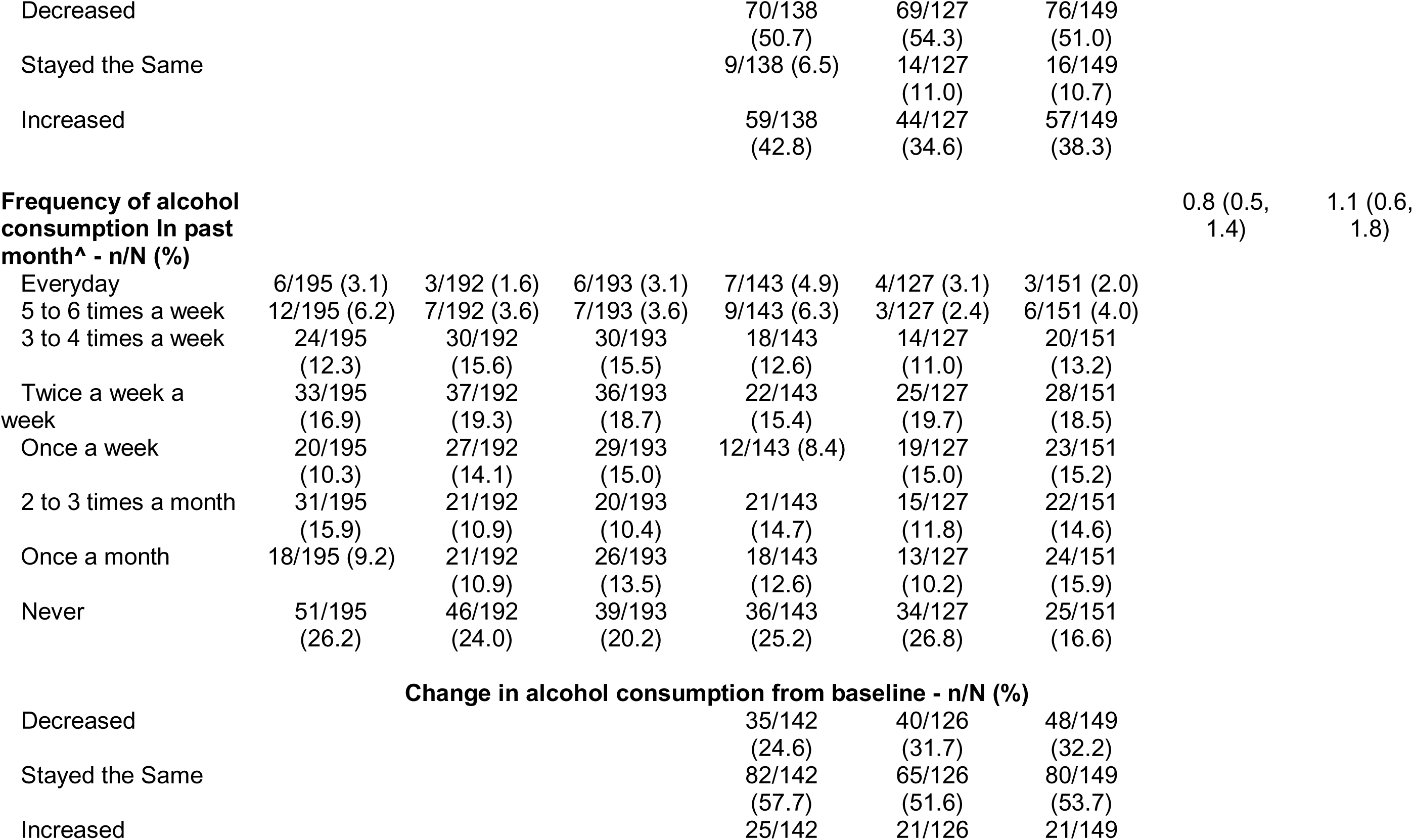
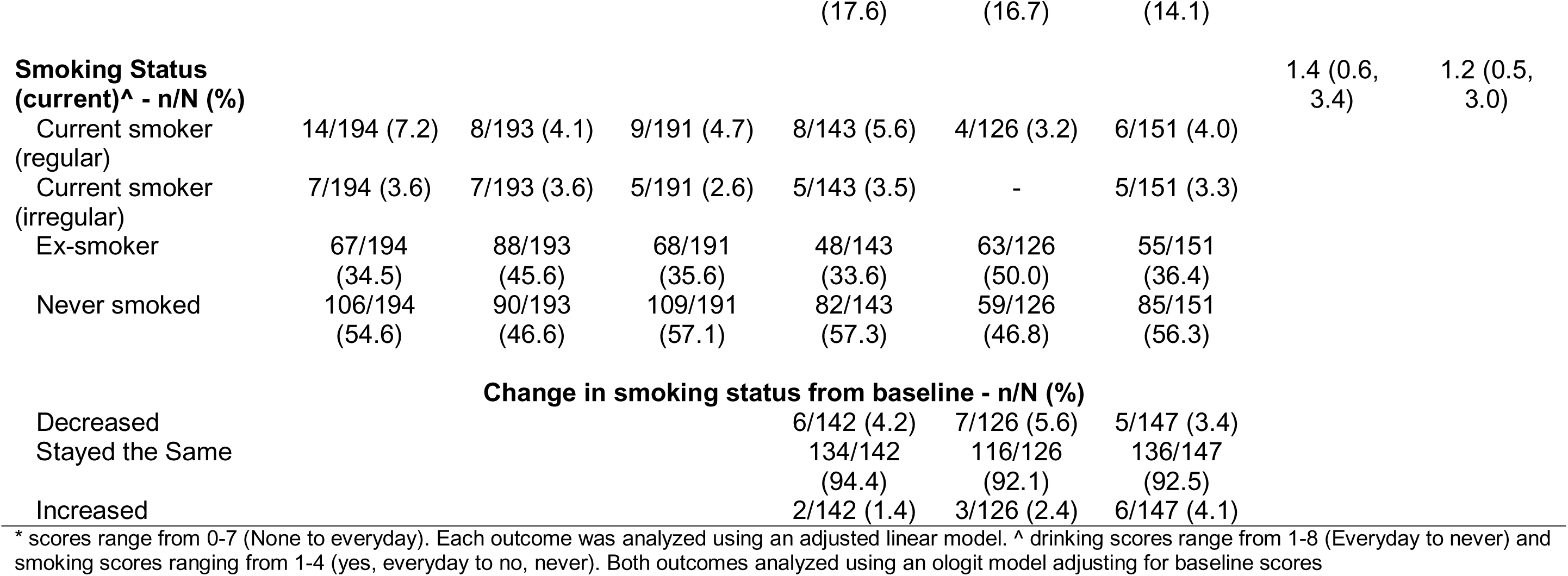
Health behaviours by treatment allocation at baseline and 12 months.

### Weight management strategies

Nine of the 15 measured weight loss strategies (see Table 4) investigated showed no differences between the texts with incentives and control groups. Compared to the control group, participants in the texts with incentives group were more likely to report self-weighing (OR 2.2 [97.5% CI 1.3, 3.5], Table 4). At 12 months, 56.8% of the texts with incentives group reported self-monitoring their weight at least once a week, compared to 37.7% in the control group. There was no difference in self-weighing between the texts alone and control groups. Moreover, there was no difference in self-monitoring pedometer steps between the control group and either the texts with incentives or texts alone groups. Compared to the control group, participants in the texts with incentives group were more likely to report avoiding certain foods (OR 3.0 (97.5% CI 1.6, 5.7]), having a weight goal to work towards (OR 4.7 [97.5% CI 2.6, 8.5]), reminding oneself of the reasons for trying to lose weight (OR 3.2 [97.5% 1.8, 5.8]), swapping one type of food for another (OR 2.1 [97.5% CI 1.2, 3.6]), and telling others about weight loss goals (OR 3.9 [97.5% CI 2.2, 7.1]). At 12 months, 65.5% of the texts with incentives group participants reported working towards a weight loss goal, compared to 34.6% of control group participants.

**Table 4:**
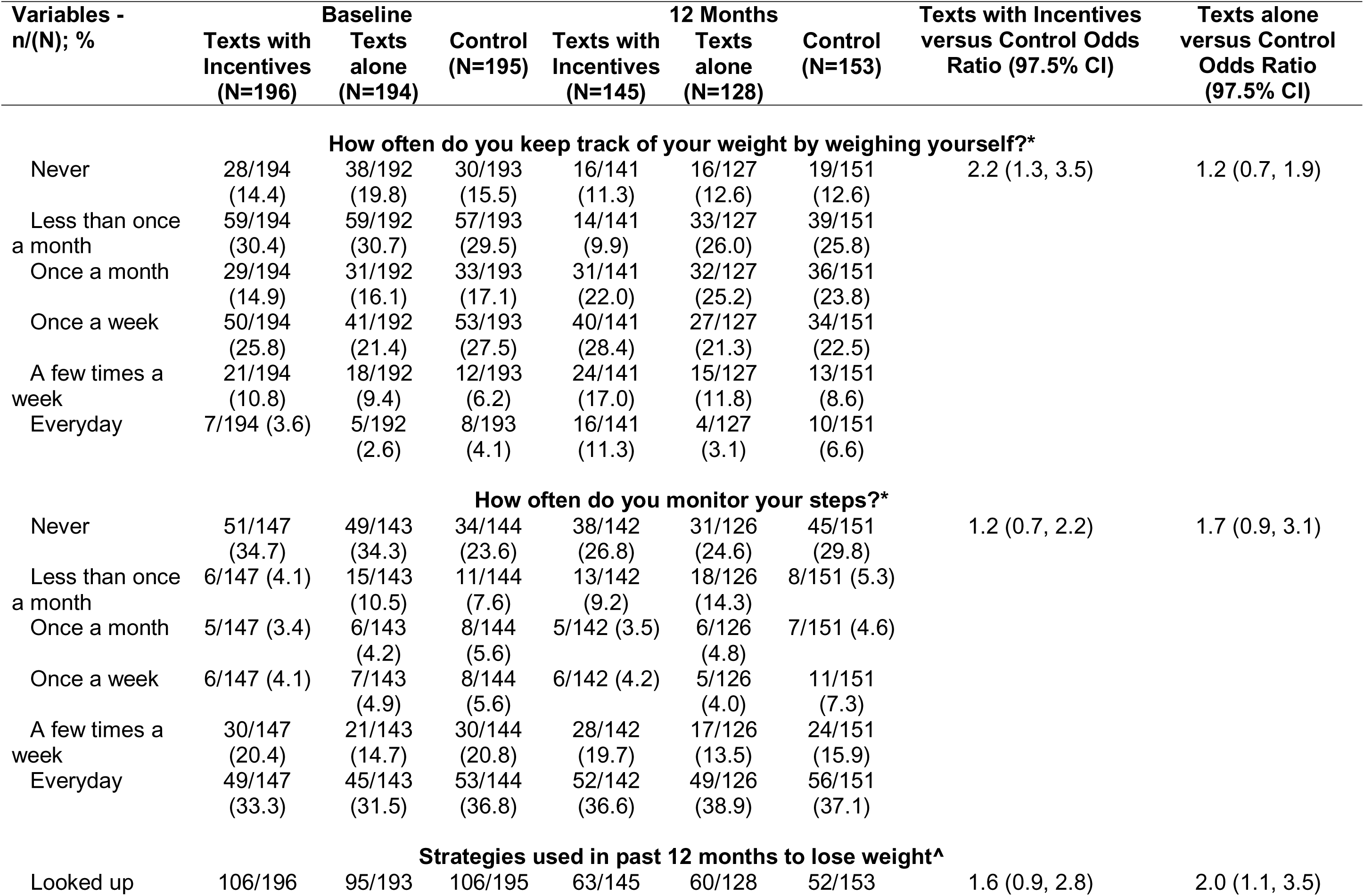
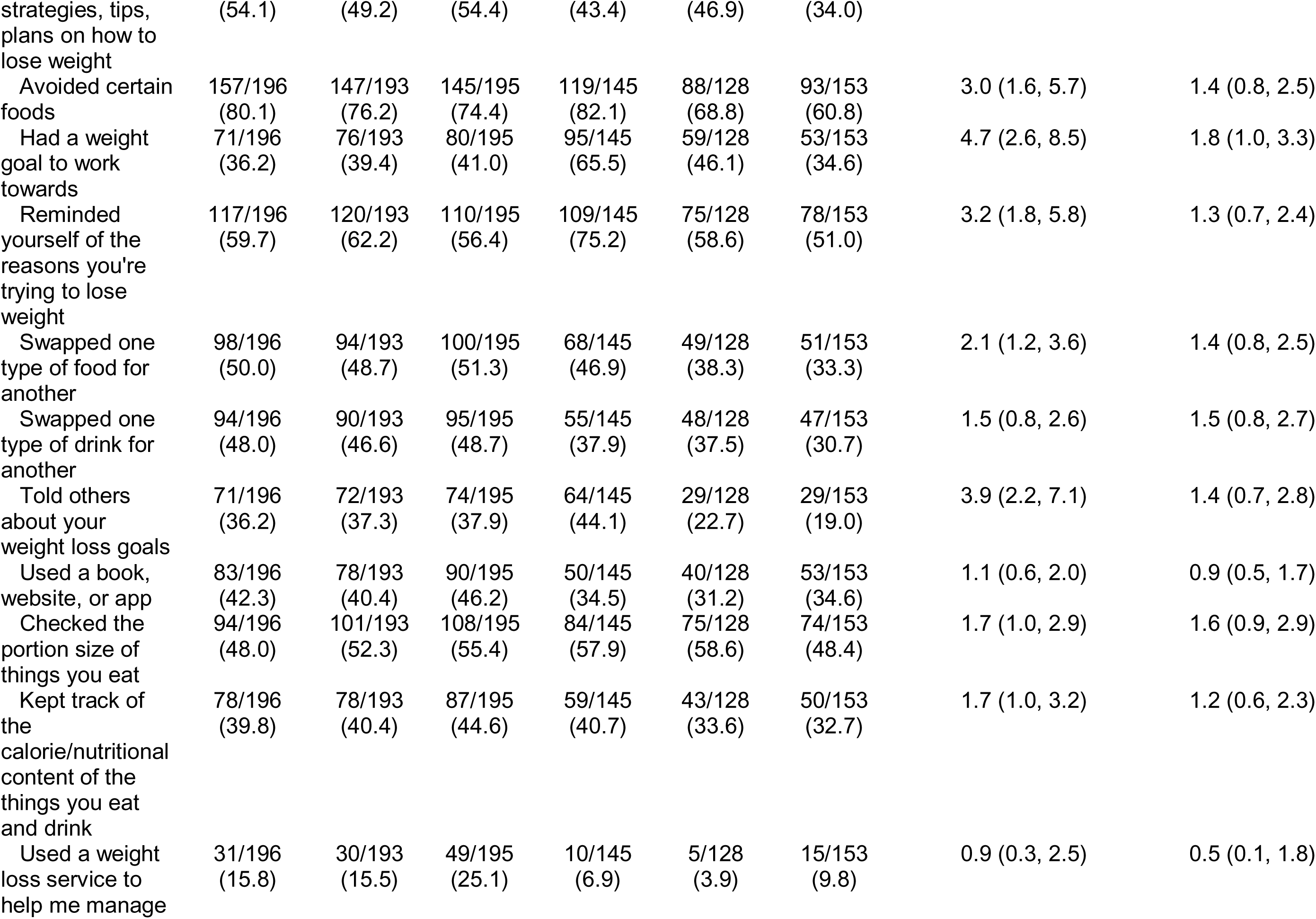
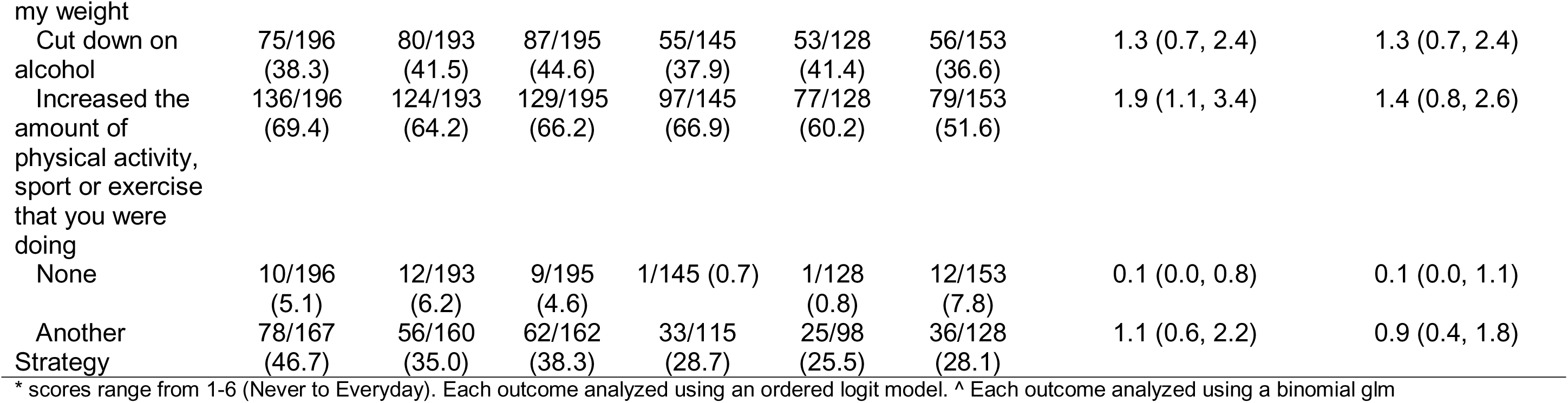
Weight management strategies by treatment allocation at baseline and 12 months.

Fourteen of the 15 weight loss strategies investigated showed no differences between the texts alone and control groups (see Table 4). Compared to the control group, participants in the texts alone group were more likely to report looking up strategies, tips, and plans on how to lose weight (OR 2.0 [97.5% CI 1.1, 3.5]).

### Weight management-related confidence

Compared to the control group, participants in the texts with incentives group had higher levels of confidence in their ability to lose weight (MD = 0.6 [97.5% CI 0.2, 1.0]) and maintain weight loss (MD = 0.9 [97.5% CI 0.5, 1.3], see Table 5). There were no differences in confidence for weight loss and weight loss maintenance between the texts alone and control group.

**Table 5:**
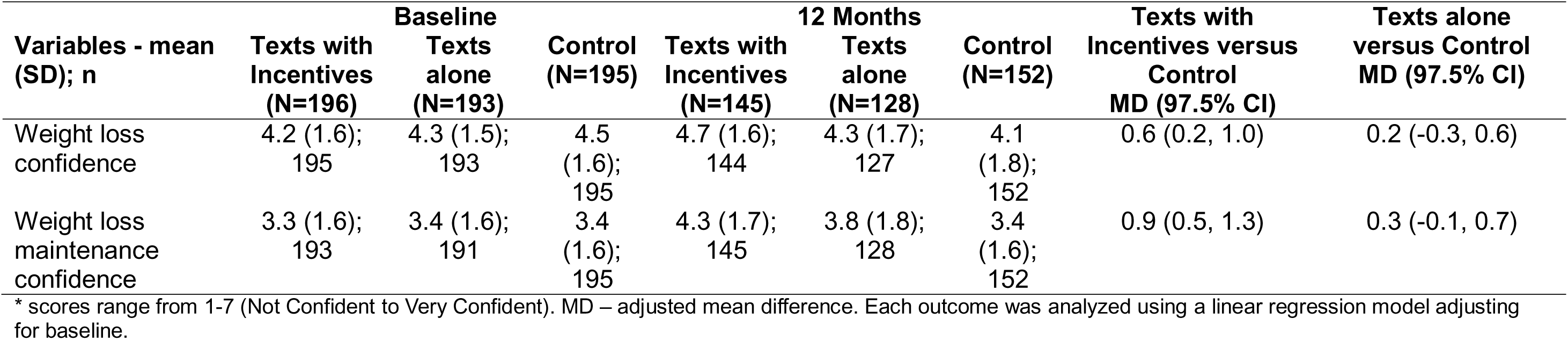
Confidence in weight management abilities by treatment allocation at baseline and 12 months.

## Discussion

This secondary exploratory analysis found little evidence of any clinically important socio-economic, health or behavioural moderators of effectiveness for the intervention effects. Hence, Game of Stones appears to be equally effective across a variety of different sub-populations within the trial when examining a multitude of pre-specified factors which have been associated with obesity. Based on these findings, the Game of Stones trial interventions of either behaviour-focused text messages alongside financial incentives, or text messages alone, are unlikely to contribute to intervention generated inequalities (7). This finding is in line with a systematic review examining inequalities in the uptake of, adherence to, and effectiveness of behavioural weight management interventions in adults, which found that most trials did not display an inequalities’ gradient (6). However, it should be noted that most trials in this systematic review were unlikely to have sufficient statistical power to identify if inequalities were present.

There was evidence of participants engaging in several evidence-based weight management strategies, particularly in the texts with financial incentives group. Engagement in strategies to facilitate behaviour change is critical for the long-term maintenance of behaviour change and weight(28). Participants in the texts with financial incentives group reported engaging in more weight management strategies, including motivational (e.g. reminding oneself of the reasons for trying to lose weight) and action focused strategies (e.g. swapping one type of food for another). Of note is that participants provided with financial incentives reported more goal setting strategies compared to control participants. All participants in this study were provided with a personalised weight loss goal following baseline measures by calculating the weight loss required for 5% and 10% weight loss, and it appears that the provision of financial incentives may have increased the relevance of the goal. Moreover, participants in the texts with incentives group reported higher levels of weight management confidence compared to the control group, suggesting that incentives alongside behaviour-focused text messages might activate a variety of psychological processes in addition to merely increasing motivation.

There was no evidence of significant changes in self-reported health-related behaviours of physical activity, sedentary behaviour, alcohol consumption or smoking status, which can affect weight loss. The lack of significant change in physical activity at both the moderate and vigorous level is unexpected, given that some evidence suggests that men often value the use of activity related behaviours for weight management (29, 30). Website information and text messages highlighted that dietary change is required to lose weight, but participants could choose the behavioural focus most relevant for them. Of relevance, a relatively high proportion of participants reported living with multiple long-term conditions and/or a disability compared to other studies, and these condition may pose additional challenges for physical activity and attending health promotion services.

The current study has several strengths. The comprehensive moderator analyses undertaken were all pre-specified and focused on several relevant factors which might potentially explain differential effects in important subgroups. Moreover, the sample recruited to this trial represents an underserved population of men, displaying high levels of obesity, socio-economic disadvantage and obesity related co-morbidities.

This study has some limitations. The sample size considerations for this study are based on changes in the primary outcome weight change at 12 months only, and the current analyses were not considered. This exploratory study presents multiple subgroup and exploratory analyses increasing the chance of type I errors. However, we have some confidence in the largely null findings since the confidence intervals suggest that we are not likely to be missing a clinically important effect size difference between the compared subgroups. Some of the subgroup classifications might have not been optimal, particularly when categorising continuous variables. Several subgroup analyses (e.g. EQ5D or alcohol consumption) had imbalances between the groups. The behavioural measures obtained were all self-reported and brief to reduce measurement burden and boost study retention, and dietary intake was not measured.

## Conclusion

The Game of Stones trial suggests equitable effectiveness for all men living with obesity regardless of socio-economic, health or wellbeing status. The texts with financial incentives group showed greater engagement in some weight management strategies and favourable changes in weight management confidence.

## Data Availability

All data produced in the present study are available upon reasonable request to the authors

## Acknowledgements

We thank all the trial participants, general practitioners and supporters who made this research possible. We also thank the following: Kathryn Machray, PhD, and Norelle Calder-McPhee, MSc, University of Stirling; Clare Jess, LLB, Christina O’Neill BSc(Hons), Angela Mullan, HND, Queens University Belfast; Hilary Taylor, MSc, Jack Brazier MSc, and all the students who assisted; Matthew McDonald, PhD, Curtin University, Australia, and other team members from the feasibility trial who shared their experiences of recruiting and collecting data; the Men’s Health Forum in Great Britian and Ireland since 2010 for invaluable contributions to the design and conduct of this study; Martin Tod, BSc(Hons) and Jim Pollard, MA, Men’s Health Forum, London; Colin Fowler, BA(Hons), Men’s Health Forum in Ireland, Dublin, Ireland; Paula Caroll, PhD, South East Technological University, Waterford, Ireland; Michael McKeon, MEd, Dublin City University, Ireland; the Trial Steering Committee: Edmund Juszczak, MSc, University of Nottingham; Emma Frew, PhD, and Kate Jolly, PhD, University of Birmingham; Graham Jameson (lay member and participant in the Football Fans in Training trial); David Gardner (lay member and Chairman of Scottish Men’s Sheds), the Scottish Men’s Sheds Association, Banchory, for their oversight and guidance; the trial protocol contributors who are no longer involved with the study: Andrew Elders, MSc, Glasgow Caledonian University, and Beatriz Goulao, PhD, Centre for Healthcare Randomised Trials, University of Aberdeen for statistics contributions; Mark Forrest, BSc, Connor Keegan, BSc(Hons), and the team at the Centre for Healthcare Randomised Trials Centre for Healthcare Randomised Trials, University of Aberdeen, for technical administrative support and database and website development; Fiona M. Harris, PhD, University of the West of Scotland, for process evaluation contributions; Claire Jones, PhD, Jack Gilmore, BSc, Ross Teviotdale, BSc(Hons), and Keith Milburn, BSc(Hons), Health Informatics Centre, University of Dundee, who developed the participant tracker software and delivered the text intervention. Finally, we acknowledge the earlier work of I. K. Crombie, MSc, University of Dundee. Mr Gardner and Mr Graham received compensation for their contributions.

## Footnotes

1 For subgroup analyses examining change in financial strain, perceived wealth, perceived enough money, and perceived wealth compared to neighbourhood see online appendix 1.

2 For subgroup analyses examining change in social weight loss reported by participants at 12 months see online appendix 1.

## References

1. GBD Obesity Collaborators. Health effects of overweight and obesity in 195 countries over 25 years. New England Journal of Medicine 2017;377: 13–27.

2. Office for Health Improvement and Disparities. Obesity Profile: short statistical commentary May 2023 2023 [cited 2024 02/20]. Available from: https://www.gov.uk/government/statistics/obesity-profile-update-may-2023/obesity-profile-short-statistical-commentary-may-2023.

3. National Institute of Diabetes and Digestive and Kidney Disease. Overweight & Obesity Statistics 2021 [cited 2024 10/02]. Available from: https://www.niddk.nih.gov/health-information/health-statistics/overweight-obesity.

4. Robertson C, Archibald D, Avenell A, Douglas F, Hoddinott P, van Teijlingen E, et al. Systematic reviews of and integrated report on the quantitative, qualitative and economic evidence base for the management of obesity in men. Health Technology Assessessment 2014;18: v-vi, xxiii-xxix, 1–424.

5. McDonald MD, Dombrowski SU, Skinner R, Calveley E, Carroll P, Elders A, et al. Recruiting men from across the socioeconomic spectrum via GP registers and community outreach to a weight management feasibility randomised controlled trial. BMC Medical Research Methodology 2020;20: 249.

6. Birch JM, Jones RA, Mueller J, McDonald MD, Richards R, Kelly MP, et al. A systematic review of inequalities in the uptake of, adherence to, and effectiveness of behavioral weight management interventions in adults. Obesity Review 2022;23: e13438.

7. Lorenc T, Petticrew M, Welch V, Tugwell P. What types of interventions generate inequalities? Evidence from systematic reviews. Journal of Epidemiology and Community Health 2013;67: 190–193.

8. Macaulay L, O’Dolan C, Avenell A, Carroll P, Cotton S, Dombrowski S, et al. Effectiveness and cost-effectiveness of text messages with or without endowment incentives for weight management in men with obesity (Game of Stones): study protocol for a randomised controlled trial. Trials 2022;23: 582.

9. Hoddinott P, O’Dolan C, Macaulay L, Dombrowski SU, Swingler J, Cotton S, et al. Text Messages With Financial Incentives for Men With Obesity: A Randomized Clinical Trial. JAMA 2024;332: 31–40.

10. Dombrowski SU, McDonald M, Van Der Pol M, Grindle M, Avenell A, Carroll P, et al. Game of Stones: feasibility randomised controlled trial of how to engage men with obesity in text message and incentive interventions for weight loss. BMJ Open 2020;10: e032653.

11. Mackenzie RM, Ells LJ, Simpson SA, Logue J. Core outcome set for behavioural weight management interventions for adults with overweight and obesity: Standardised reporting of lifestyle weight management interventions to aid evaluation (STAR-LITE). Obesity Review 2020;21: e12961.

12. O’Neill J, Tabish H, Welch V, Petticrew M, Pottie K, Clarke M, et al. Applying an equity lens to interventions: using PROGRESS ensures consideration of socially stratifying factors to illuminate inequities in health. Journal of Clinical Epidemiology 2014;67: 56–64.

13. Welch VA, Norheim OF, Jull J, Cookson R, Sommerfelt H, Tugwell P. CONSORT-Equity 2017 extension and elaboration for better reporting of health equity in randomised trials. BMJ 2017;**359**: j5085.

14. Petkovic J, Duench SL, Welch V, Rader T, Jennings A, Forster AJ, et al. Potential harms associated with routine collection of patient sociodemographic information: A rapid review. Health Expectations 2019;22: 114–129.

15. Abel GA, Barclay ME, Payne RA. Adjusted indices of multiple deprivation to enable comparisons within and between constituent countries of the UK including an illustration using mortality rates. BMJ Open 2016;6: e012750.

16. Office for National Statistics. GSS harmonisation support 2020 [cited 2024 08/06]. Available from: https://analysisfunction.civilservice.gov.uk/government-statistical-service-and-statistician-group/gss-support/gss-harmonisation-support/.

17. Scottish Government. Scottish Surveys: Core and Harmonised Questions 2017 [cited 2024 08/06]. Available from: https://www.gov.scot/publications/scottish-surveys-core-and-harmonised-questions/.

18. Best M, Papies EK. Lower socioeconomic status is associated with higher intended consumption from oversized portions of unhealthy food. Appetite 2019;140: 255–268.

19. French D. Financial strain in the United Kingdom. Oxford Economic Papers 2017;70: 163–182.

20. Herdman M, Gudex C, Lloyd A, Janssen M, Kind P, Parkin D, et al. Development and preliminary testing of the new five-level version of EQ-5D (EQ-5D-5L). Quality of Life Research 2011;20: 1727–1736.

21. Tennant R, Hiller L, Fishwick R, Platt S, Joseph S, Weich S, et al. The Warwick-Edinburgh Mental Well-being Scale (WEMWBS): development and UK validation. Health and Quality of Life Outcomes 2007;5: 63.

22. Kroenke K, Spitzer RL, Williams JB, Löwe B. An ultra-brief screening scale for anxiety and depression: the PHQ-4. Psychosomatics 2009;50: 613–621.

23. Lillis J, Luoma JB, Levin ME, Hayes SC. Measuring weight self-stigma: the weight self-stigma questionnaire. Obesity 2010;18: 971–976.

24. Office for National Statistics. Disability variable: Census 2021 2021 [cited 2024 10/17]. Available from: https://www.ons.gov.uk/census/census2021dictionary/variablesbytopic/healthdisabilityandunpaidcarevariablescensus2021/disability.

25. Craig CL, Marshall AL, Sjöström M, Bauman AE, Booth ML, Ainsworth BE, et al. International physical activity questionnaire: 12-country reliability and validity. Medicine & Science in Sports & Exercise 2003;35: 1381–1395.

26. Hartmann-Boyce J, Aveyard P, Piernas C, Koshiaris C, Velardo C, Salvi D, et al. Cognitive and behavioural strategies for weight management in overweight adults: Results from the Oxford Food and Activity Behaviours (OxFAB) cohort study. PLoS One 2018;13: e0202072.

27. LeBlanc ES, Patnode CD, Webber EM, Redmond N, Rushkin M, O’Connor EA. Behavioral and Pharmacotherapy Weight Loss Interventions to Prevent Obesity-Related Morbidity and Mortality in Adults: Updated Evidence Report and Systematic Review for the US Preventive Services Task Force. JAMA 2018;320: 1172–1191.

28. Hankonen N. Participants’ enactment of behavior change techniques: a call for increased focus on what people do to manage their motivation and behavior. Health Psychology Review 2021;15: 185–194.

29. Hunt K, McCann C, Gray CM, Mutrie N, Wyke S. “You’ve got to walk before you run”: positive evaluations of a walking program as part of a gender-sensitized, weight-management program delivered to men through professional football clubs. Health Psychology 2013;32: 57–65.

30. Archibald D, Douglas F, Hoddinott P, van Teijlingen E, Stewart F, Robertson C, et al. A qualitative evidence synthesis on the management of male obesity. BMJ Open 2015;5: e008372.

