## Supplementary file for "Secondary analysis of the Game of Stones trial of text messages with financial incentives for men with obesity"

**Online supplement 1 - Moderator subgroup analyses: Weight change % at 12 months in relation to changes in perceived wealth and financial strain.**

| **Subgroup*** | **Texts with Incentives (n=146)** | **Texts alone (n=128)** | **Control (n=152)** | **Texts with Incentives versus Control Interaction Effect: MD (99.5% CI); p value** | **Texts alone versus Control**  **Interaction Effect: MD (99.5% CI); p value** |
| --- | --- | --- | --- | --- | --- |
| **Change in Financial Strain** | | | | | |
| Decrease | -6.8 (9.3); 4 | -5.6 (9.5); 5 | -4.5 (7.2); 3 |  |  |
| Stayed The Same | -4.9 (6.1); 122 | -2.9 (6.3); 103 | -1.0 (5.4); 129 | -1.65 (-14.74, 11.45); 0.72 | -0.73 (-13.30, 11.83); 0.87 |
| Increase | -2.3 (4.1); 5 | 1.8 (3.9); 8 | -3.7 (6.7); 8 | 3.12 (-13.05, 19.28); 0.59 | 6.50 (-8.51, 21.51); 0.22 |
| **Change in Perceived Enough Money** | | | | | |
| Decrease | -4.8 (4.5); 24 | -3.0 (8.3); 32 | -2.2 (7.5); 34 |  |  |
| Stayed The Same | -4.8 (6.3); 81 | -2.7 (6.1); 68 | -0.1 (4.2); 76 | -1.88 (-7.19, 3.44); 0.32 | -1.63 (-6.74, 3.47); 0.37 |
| Increase | -5.2 (7.0); 36 | -2.7 (3.8); 17 | -1.5 (5.3); 31 | -1.03 (-7.22, 5.16); 0.64 | -0.49 (-7.21, 6.22); 0.84 |
| **Change in Perceived Wealth** | | | | | |
| Decrease | -5.3 (5.2); 16 | -1.6 (5.5); 24 | -2.3 (7.6); 31 |  |  |
| Stayed The Same | -4.7 (6.2); 81 | -3.3 (7.8); 61 | -0.4 (4.5); 72 | -1.38 (-7.38, 4.62); 0.52 | -3.76 (-9.33, 1.81); 0.06 |
| Increase | -6.0 (6.4); 39 | -2.6 (3.8); 28 | -1.0 (5.2); 34 | -2.13 (-8.81, 4.54); 0.37 | -2.64 (-9.11, 3.84); 0.25 |
| **Change in Perceived Wealth compared to Neighbourhood** | | | | | |
| Decrease | -4.9 (7.4); 16 | 0.1 (4.3); 22 | -1.4 (7.7); 36 |  |  |
| Stayed The Same | -5.3 (5.8); 87 | -3.6 (7.5); 68 | -0.7 (4.9); 73 | -1.05 (-6.93, 4.83); 0.61 | -4.41 (-9.94, 1.12); 0.03 |
| Increase | -4.6 (7.1); 32 | -3.0 (4.3); 24 | -1.5 (3.9); 32 | 0.24 (-6.55, 7.02); 0.92 | -3.18 (-9.83, 3.47); 0.18 |

**Online supplement 2 - Moderator subgroup analyses: Percent weight change at 12 months from baseline in relation to for social weight loss reported by participants at 12 months**

|  | **Texts with incentives (n=146)** | **Texts alone (n=128)** | **Control (n=152)** | **Texts with Incentives Interaction Effect: MD (99.5% CI); p value** | **Texts alone Interaction Effect: MD (99.5% CI); p value** |
| --- | --- | --- | --- | --- | --- |
| **Who have you told about your participation in Game of Stones** | | | | | |
| Told no-one about Game of Stones | -1.9 (6.3); 4 | -1.5 (3.5); 10 | -0.7 (4.7); 24 |  |  |
| Told others about participating in Game of Stones | -5.0 (6.1); 140 | -2.8 (6.5); 118 | -1.4 (5.6); 128 | -2.30 (-11.69, 7.09); 0.49 | -0.67 (-7.39, 6.05); 0.78 |
| **Social Weight Loss Strategy** | | | | | |
| Exclusively tried to lose weight alone | -5.2 (5.9); 108 | -3.1 (6.8); 87 | -1.7 (5.2); 81 |  |  |
| Exclusively tried to lose weight with others | -8.0 (7.1); 9 | -2.8 (5.1); 13 | -0.8 (7.9); 26 | -3.72 (-10.67, 3.23); 0.13 | -0.61 (-6.88, 5.66); 0.78 |
| Sometimes alone and sometimes with others | -3.5 (6.8); 21 | -1.2 (4.3); 24 | -1.4 (5.1); 21 | 1.44 (-4.30, 7.18); 0.48 | 1.77 (-3.89, 7.42); 0.38 |
| Did not try to lose weight | 3.0 (3.3); 4 | -0.5 (1.5); 3 | -0.4 (3.0); 21 | 6.81 (-2.67, 16.29); 0.04 | 1.64 (-9.04, 12.32); 0.66 |
